# Professional grief in cancer care – A scoping review

**DOI:** 10.1101/2024.08.07.24311612

**Authors:** Svenja Wandke, Hannah Führes, Mareike Thomas, Klaus Lang, Martin Härter, Karin Oechsle, Isabelle Scholl

**Affiliations:** Center for Psychosocial Medicine, Department of Medical Psychology, University Medical Center Hamburg-Eppendorf, Hamburg, Germany; II. Department of Medicine, University Medical Center Hamburg-Eppendorf, Hamburg, Germany; Psychotherapeutic Practice, Munich, Germany; Palliative Care Unit, Department of Oncology, Hematology and BMT, University Medical Center Hamburg-Eppendorf, Hamburg, Germany

**Author notes:** Correspondence to: Svenja Wandke, University Medical Center Hamburg-Eppendorf, Department of Medical Psychology, Martinistraße 52, Building W26, 20246 Hamburg, GERMANY.

**Keywords:** cancer care, healthcare professionals, grief over patient loss, professional grief, scoping review

## Abstract

**Objective:** Healthcare professionals (HCPs) in cancer care often face patient deaths, yet there is a notable absence of comprehensive evidence regarding their grief. This scoping review seeks to identify key aspects of professional grief in cancer care and give an overview pertaining its’ conceptualization and frequency and intensity.

**Methods:** The primary search covered three databases (PUBMED, PSYNDEX, and PsycINFO). Two independent reviewers assessed 2,248 records, selecting 34 eligible articles.

**Results:** Most studies originated from North America and Israel, with limited evidence from the global south, East Asia and Europe, as well as few quantitative studies. HCPs exhibit classic grief symptoms (such as sadness) and distinct features (e.g., feelings of guilt) in response to patient deaths, though a clear definition and measures of professional grief are lacking. Grief frequency varies highly (from 23% to 100%).

**Conclusions:** Future research should refine definitions and measures to better support HCPs in effectively managing professional grief in cancer care.

## Background

When individuals experience the loss of family members or friends, the typical and anticipated reaction is grief, which is the human, emotional response to the loss of someone or something significant (1–3). However, evidence suggests that this notion of significance also applies to professional relationships, such as those between healthcare professionals (HCPs) and their patients. In cancer care, HCPs often form close bonds with patients due to the extensive and sometimes long-term care they provide (4,5). As cancer remains a leading global cause of death (3,6), the loss of patients is inherent to HCPs working in this field. The distress HCPs might experience after a patient’s death is sometimes called “professional grief”, a phenomenon that hasn’t been comprehensively evaluated (7). While it has been suggested to differentiate between professional and personal grieving experiences (8), the distinction remains blurry. This lack of specificity and recognition is increasingly problematic as emerging evidence indicates that professional grief might contribute to elevated rates of burnout, compassion fatigue, and compromised healthcare quality (3,8,9). Thus, the overarching aim of this scoping review was to accumulate insights into professional grief within cancer care and provide a comprehensive overview of the existing body of evidence. The primary objective was to clarify fundamental concepts of professional grief in cancer care and ascertain its frequency and depth among oncology professionals.

## Methods

### Protocol and registration

To conduct this scoping review, we applied the Arksey & O’Malley framework and based our study protocol on the PRISMA-ScR-Checklist (10,11). We initially registered our study protocol on 15 September 2022 with the open science framework (https://doi.org/10.17605/OSF.IO/R7B8H), and then updated it on 13 March 2023 (https://doi.org/10.17605/OSF.IO/Y5QHN), to refine our inclusion criteria.

### Eligibility criteria

We included studies focusing on HCPs’ responses to patient death, coping strategies, and related aspects in cancer care, regardless of whether they used the term “professional grief”. Original research using qualitative, quantitative, or mixed-methods, as well as systematic reviews, were included. Articles had to be in English, German, or French (see supplementary file 1 for the checklist of inclusion criteria). We excluded articles on non-patient-related grief, non-cancer contexts, and studies involving HCPs still in training.

### Information sources and screening process

We conducted primary searches between June and September 2022, updating in April 2024 in PUBMED, PsycINFO, and PSYNDEX databases. Thus, all articles published between inception and April 2024 were considered. The search strategy for PUBMED is detailed in supplementary file 2. Records were screened in Rayyan, with duplicates removed (12). Reference tracking was also performed. Pre-screening was conducted to establish inclusion criteria consistency. Titles and abstracts were independently screened by two reviewers, with disagreements resolved through discussion. Full-text assessment was done by two reviewers independently (SW, HF), with disagreements resolved through discussion with a third reviewer (MT).

### Data charting process

The first author developed a data charting form, which was iteratively updated and independently charted by a second reviewer (LW, cf. acknowledgements), and checked by a third reviewer (MT), who recoded results when necessary. Disagreements were resolved through discussion. The charting included study characteristics, aims, key findings, terminology used, professional grief characteristics, and its association with related constructs. Upon charting, all eligible studies were categorized inductively based on their aim and key findings. Categories were developed by the first author and agreed upon by all authors.

## Results

### Included studies

After removing duplicates, we found 2,248 records in electronic databases and 47 through reference tracking. We screened 274 full-text reports, resulting in 34 eligible studies (see Figure 1). These studies, published between 1994 and 2023, mainly originated in Canada (n=10), the United States (n=9), and Israel (n=7). Most studies were qualitative, with only a fifth using validated quantitative measures to assess professional grief. They primarily focused on physicians (n=16) and nurses (n=11). Characteristics of these studies are detailed in Table 1.

**Figure 1.**
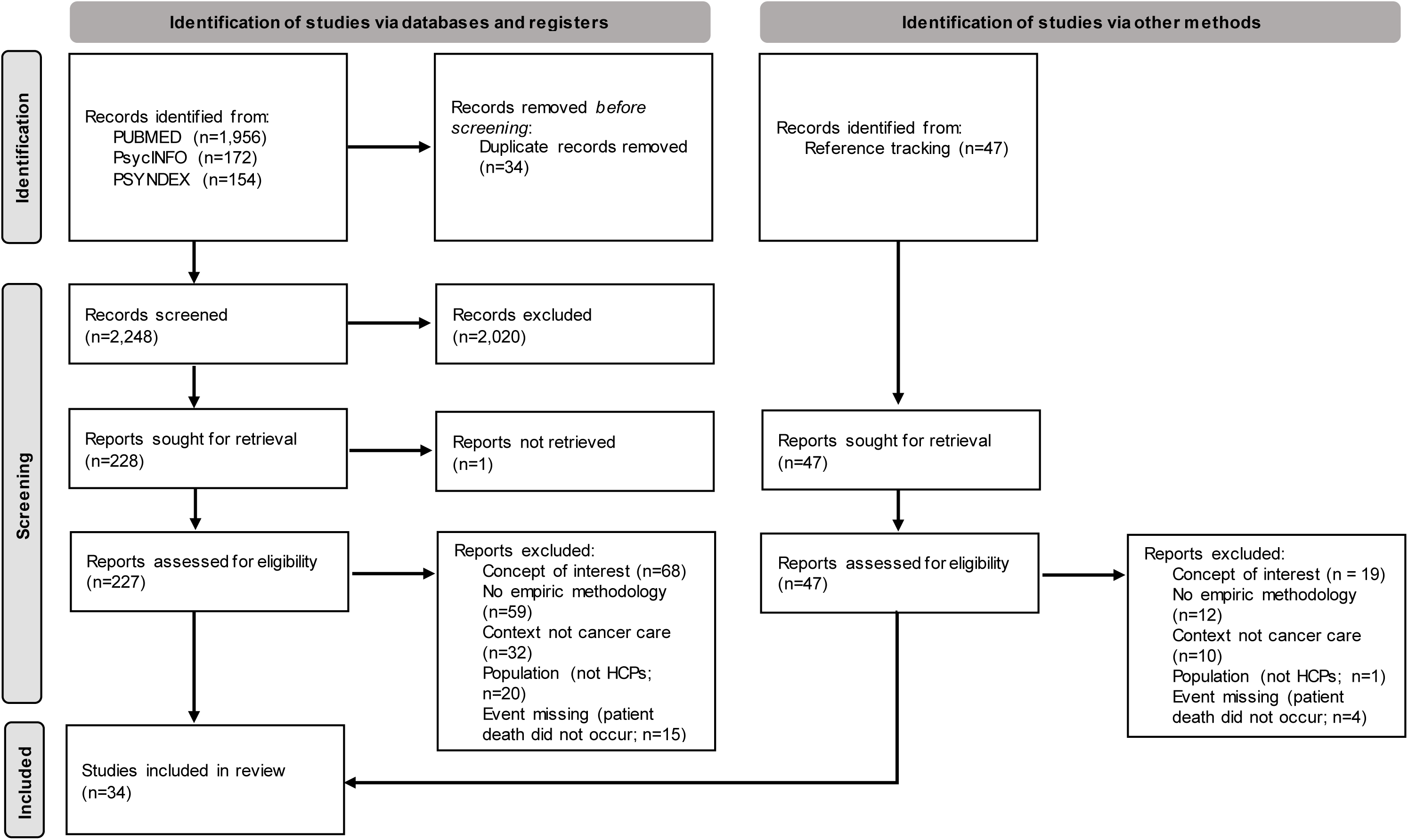
Study selection.

**Table 1.**
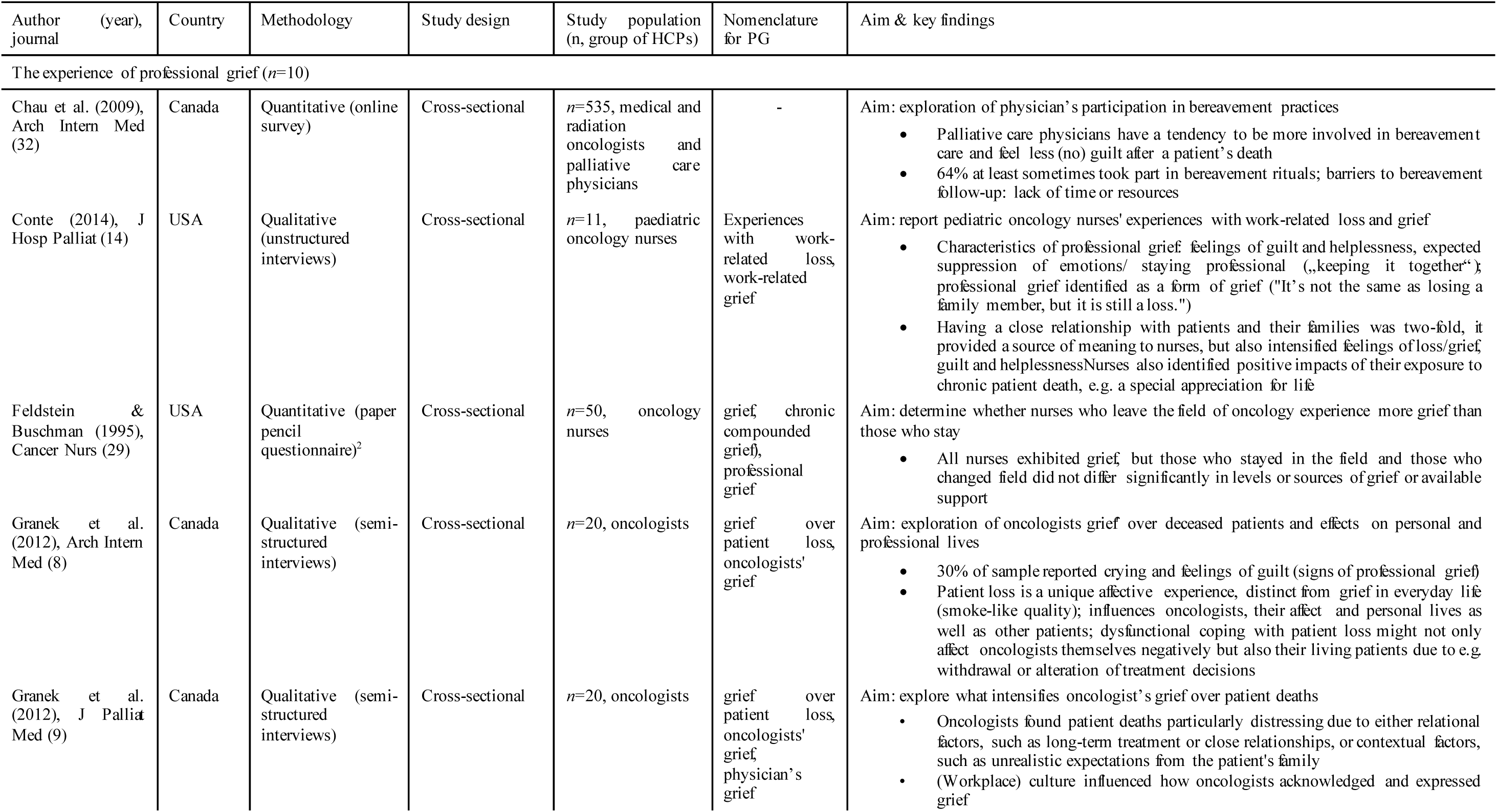
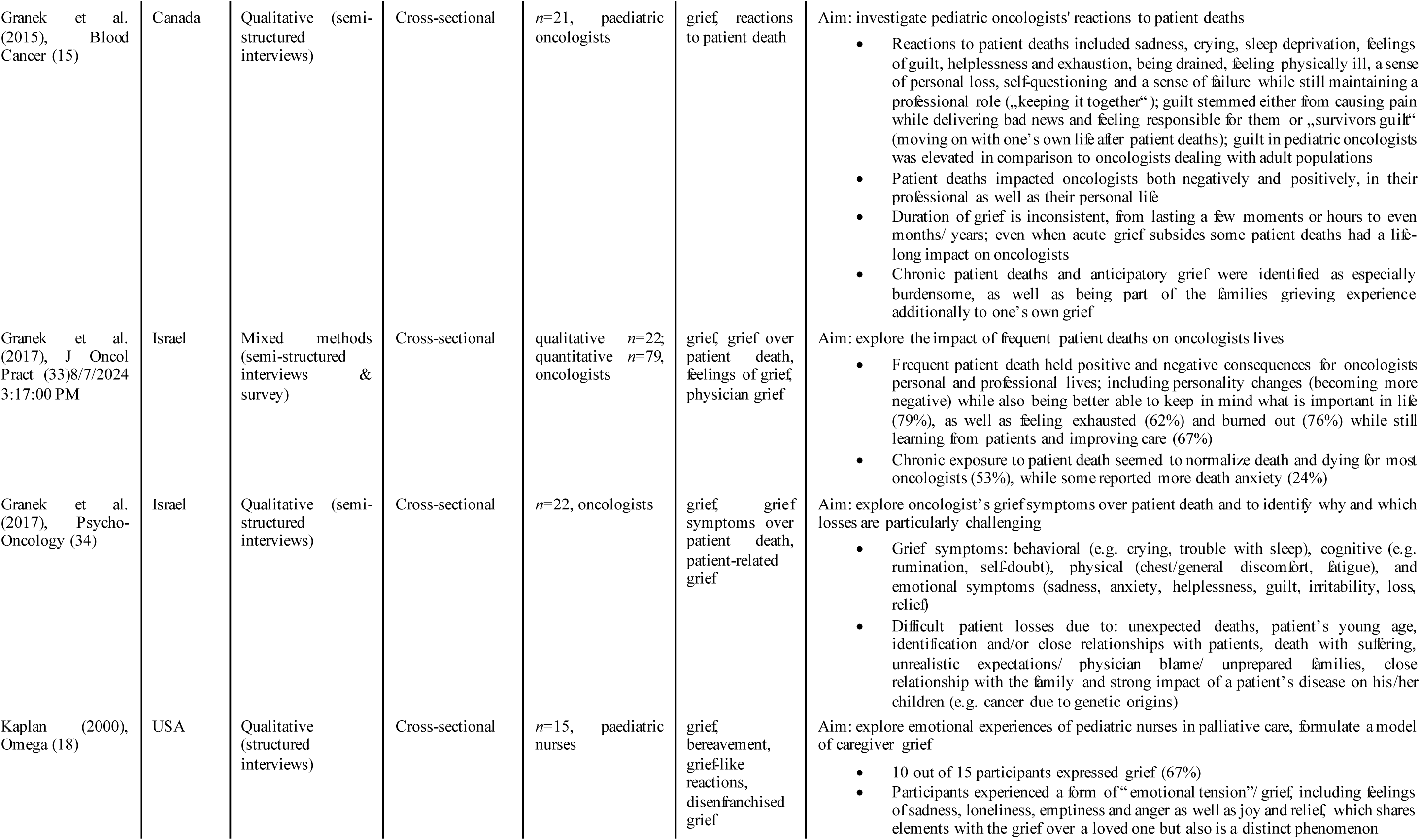
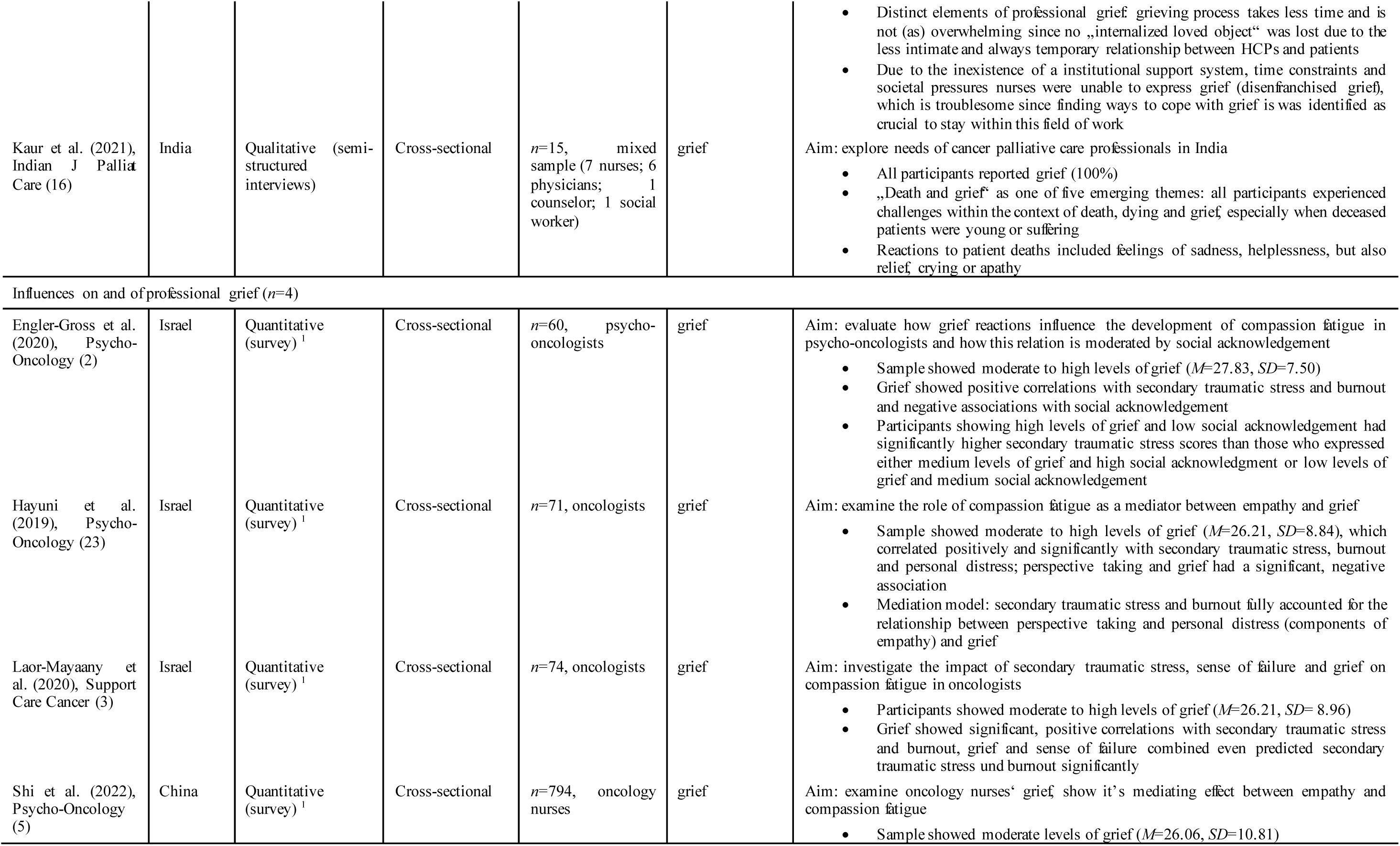
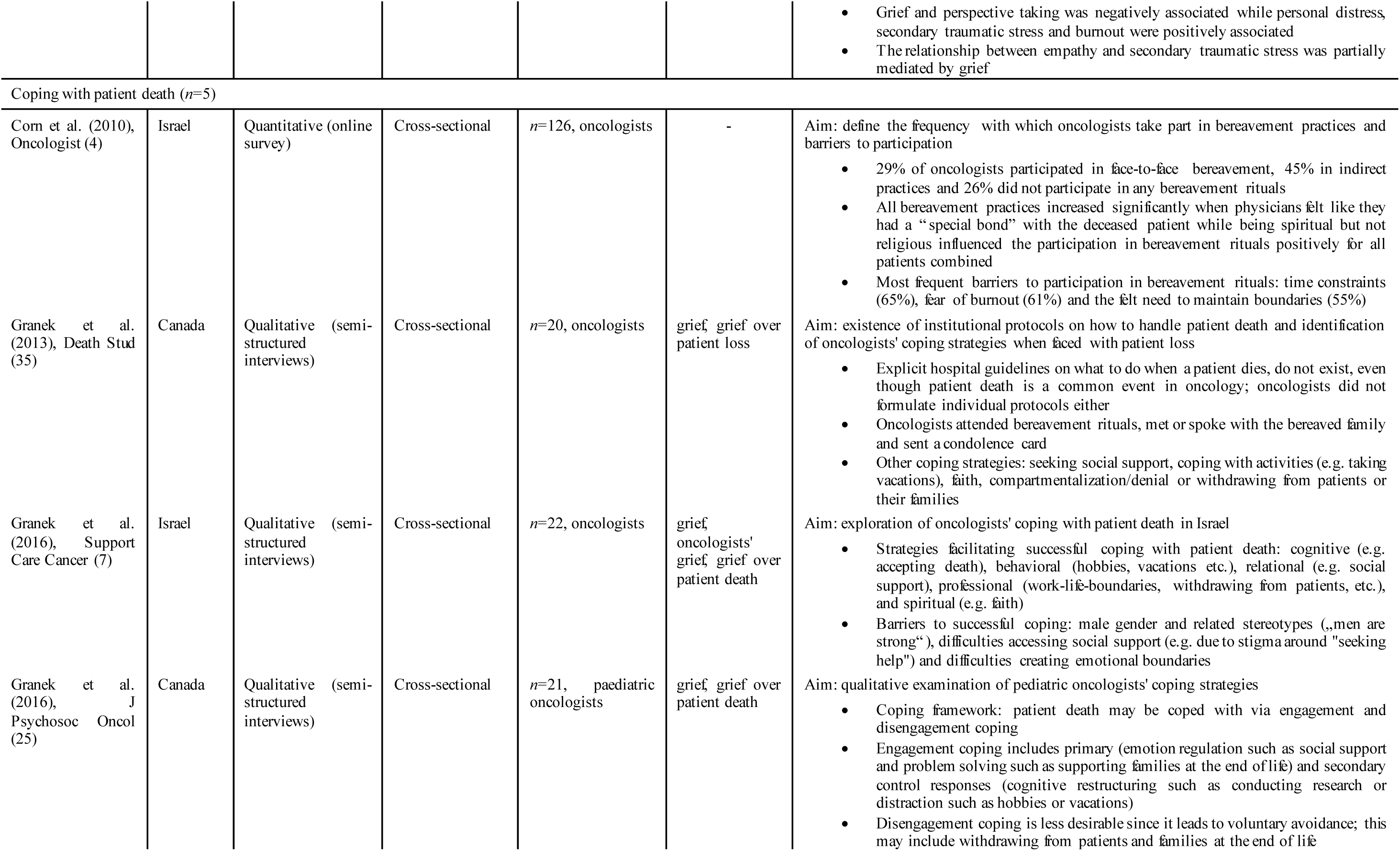
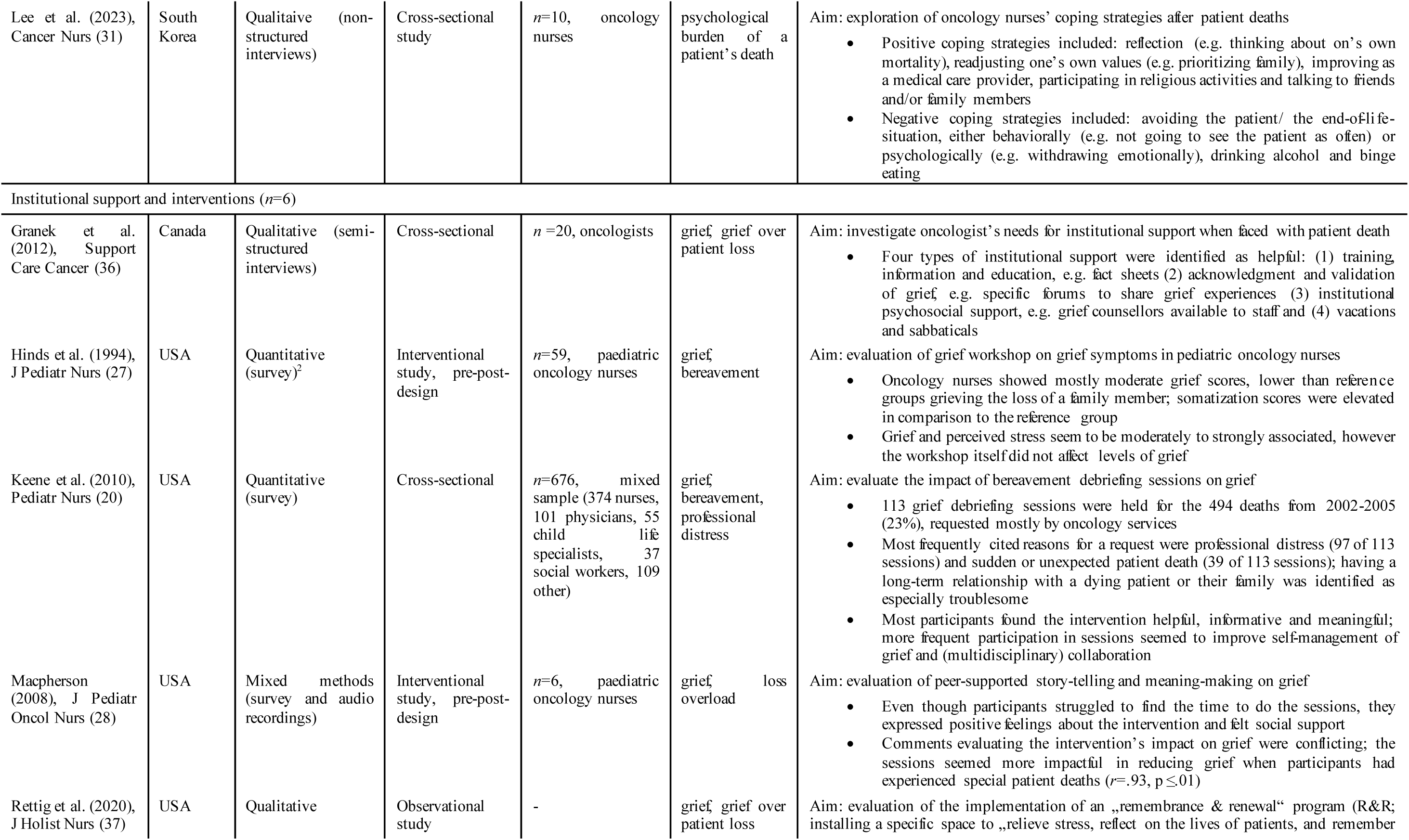
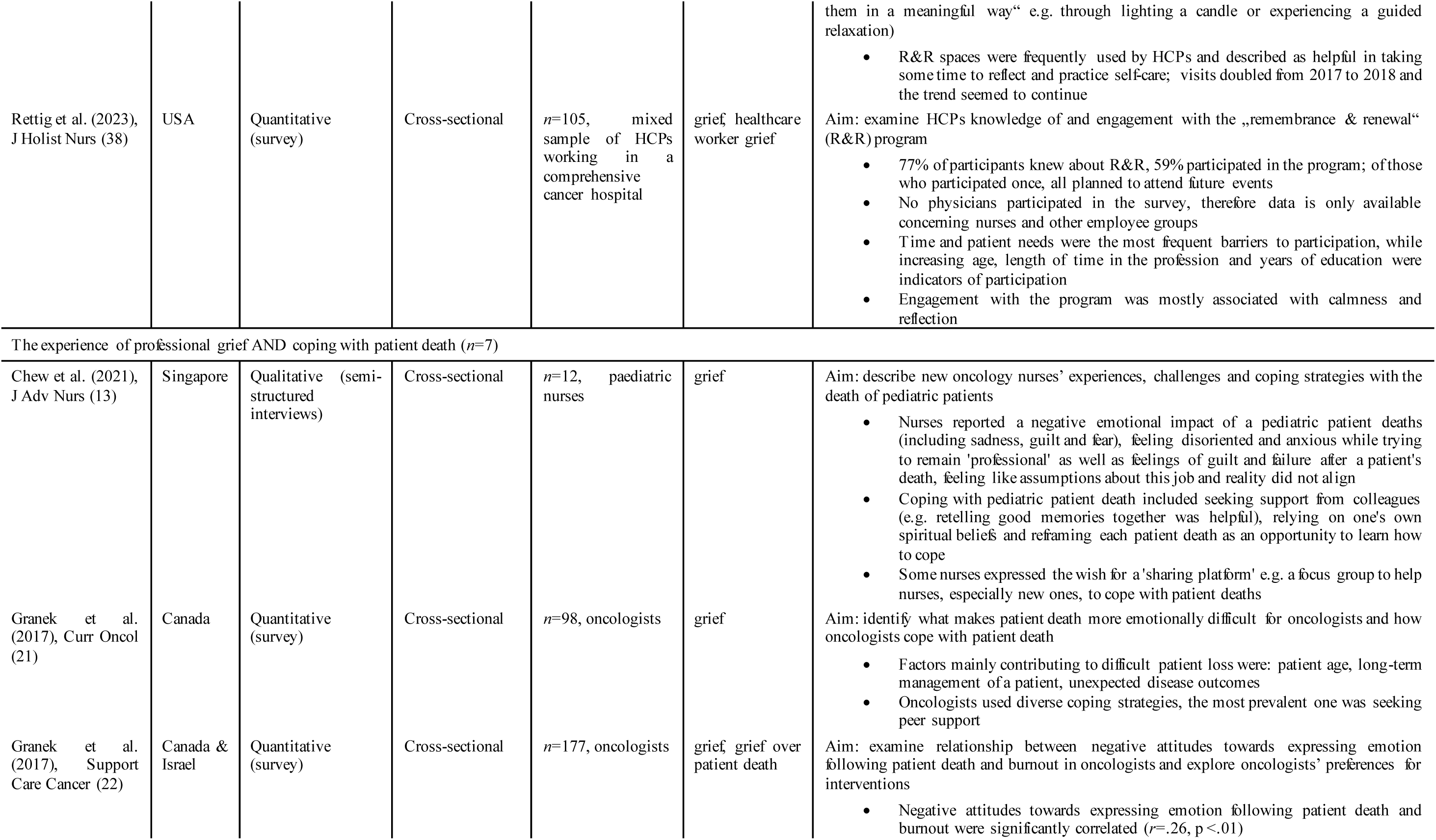
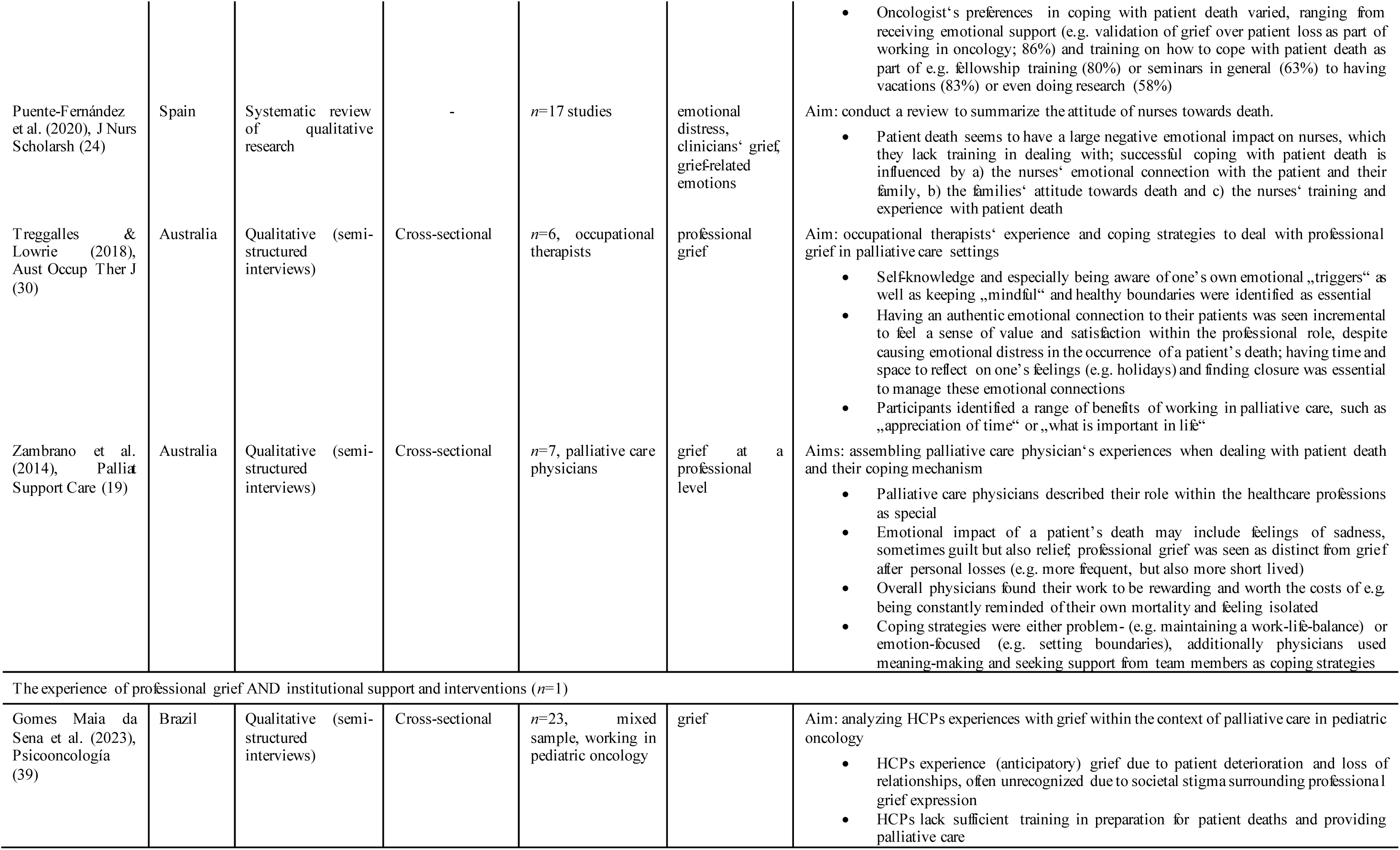
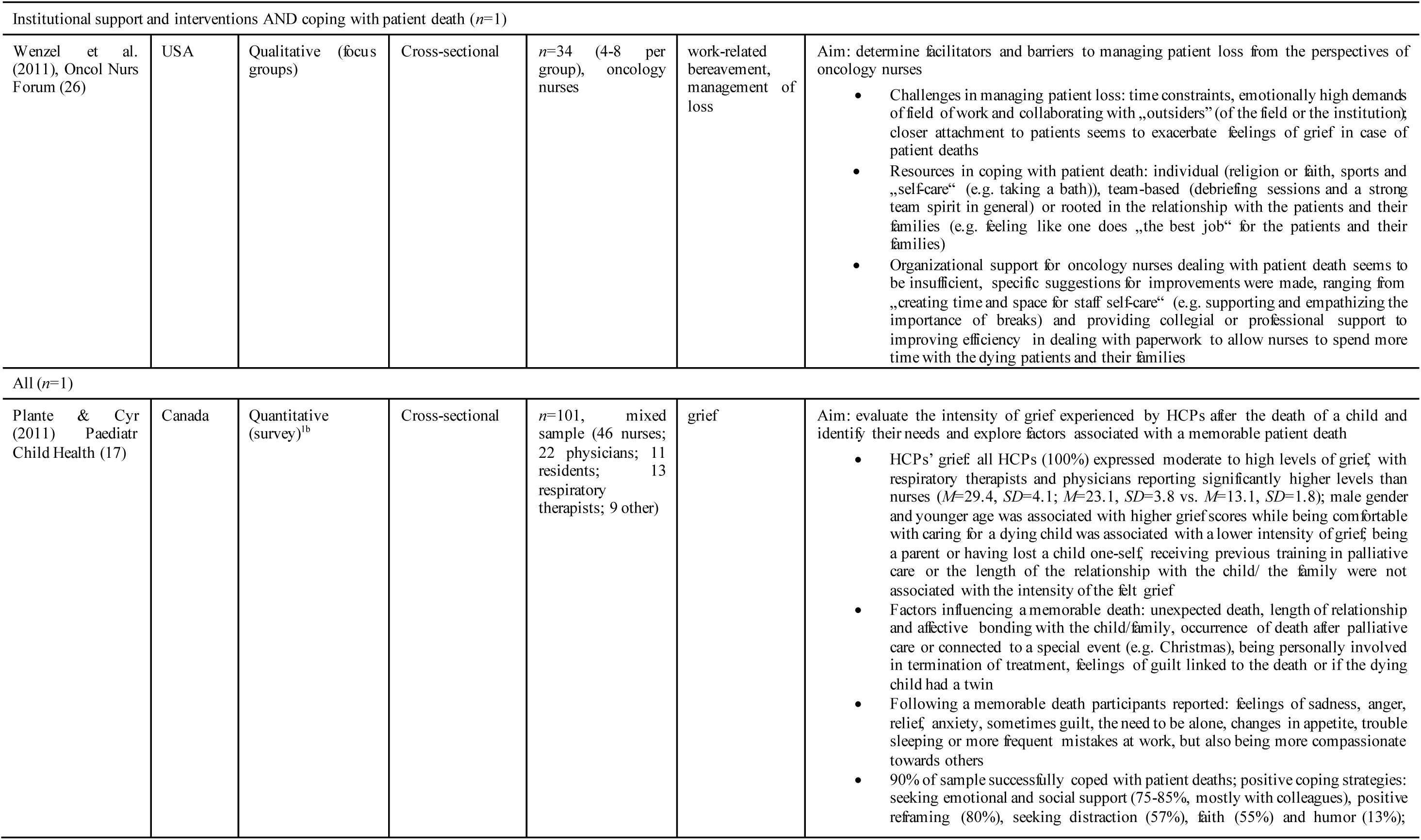
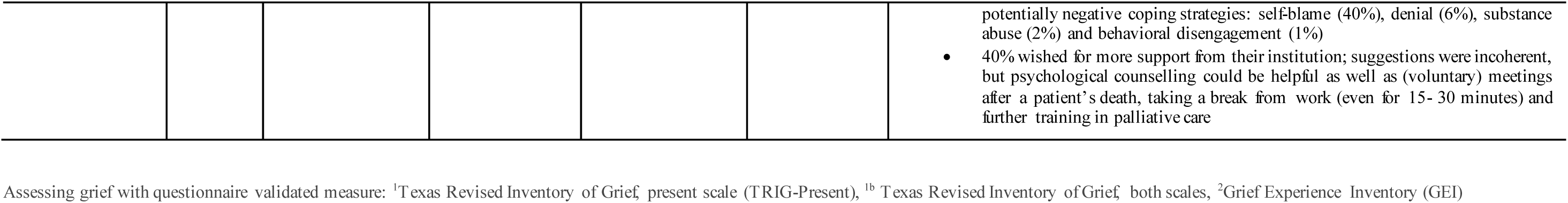
Study Characteristics.

### Synthesis of results

Data charting revealed four main categories: “the experience of professional grief”, “influences on and of professional grief”, “coping with patient death”, and “institutional support and interventions”. Some studies addressed multiple categories, with varying numbers of studies per category (1-10, see Table 1).

#### The experience of professional grief

The first category explores the emotional dimensions of professional grief, examining intensity, frequency, effects and unique contextual factors. In response to patient deaths, HCPs experience a distinctive form of grief, encompassing typical expressions like crying and sadness, along with specific aspects such as guilt and helplessness (13–17). This grief differs from personal grief due to the frequency of patient deaths in cancer care and the responsibility felt toward patients’ lives, albeit with reduced intensity (15,18,19). Studies show highly variable frequencies of professional grief, ranging from 23% to 100%, indicating multifactorial causes beyond patient deaths (8,16–18,20). Factors making patient deaths challenging include the patient’s age, closeness of the patient-HCP relationship, unexpected outcomes, and family issues (9,17,21,22). HCPs also feel pressure to maintain composure, which can lead to personality changes and exhaustion (2,5,8,23,24). However, positive outcomes include normalizing death, learning from patients, and enhancing life appreciation (21,24).

#### Influences on and of professional grief

This category explores the relationship between professional grief and other side effects of caring for terminally ill patients. Positive associations between grief and compassion fatigue and burnout were found, and grief partially mediates the link between empathy and secondary traumatic stress. Social acknowledgment of grief affects levels of secondary traumatic stress (2,3,5,23).

#### Coping with patient death

This category focuses on HCPs’ approaches to managing patient loss and the extent to which these approaches aid in effective coping. HCPs employ various coping strategies, including seeking social support, relying on faith, engaging in hobbies, and using positive reframing. Barriers to coping include time constraints, heavy workloads, and gender-related stereotypes. Some HCPs resort to detrimental strategies like emotional withdrawal or substance abuse (7,17,19,25).

#### Institutional support and interventions

Institutions can mitigate the impact of patient losses by providing training, validation of grief, opportunities for breaks, and psychosocial support (22,26). Providing or fostering collegial support emerged as beneficial (17,22). Interventions such as grief debriefing sessions and peer support workshops have proven helpful (20,27,28).

### The construct of professional grief

We assessed the existing terminology and definitions pertaining to professional grief. While the majority of articles uses terms like “grief” to depict distress following a patient’s loss, few specifically mention “professional grief” (29,30; see table 1, nomenclature). Some use rather descriptive terms such as “psychological burden of a patient’s death” or “work-related bereavement” (26,31). Although “professional grief” was already coined in 1995 (29), subsequent research did not uniformly adopt this term. Furthermore, less than 10% of articles provide definitions for professional grief (n=3; see supplementary file 3; 7,8,30). These definitions generally align with the emotional impact of professional grief outlined above but vary in focus, with Granek et al. emphasizing oncologists’ experiences (7,8), while Treggalles and Lowrie consider professional grief within the broader context of grief in general (30). Despite these efforts to define professional grief, consistent terminology and an established and comprehensive definition of the construct are lacking across the available literature.

## Discussion

In this scoping review, we analyzed 34 studies on professional grief among HCPs in cancer care, primarily from the United States, Canada, and Israel, with limited evidence from other regions. Notably, a small number of research teams provided most of the available evidence on professional grief.

Qualitative studies focusing on nurses and physicians constitute the primary source of existing knowledge, reflecting their prevalence among HCPs. However, evidence on other HCP groups like psychologists, social workers, and physical therapists is limited. Two studies suggest that psycho-oncologists and respiratory therapists also experience significant grief levels (2,17). Future research should address a broader range of occupational roles in cancer care.

Just like general grief, professional grief can be a natural and adaptive response to loss, rather than inherently pathological. Current research on professional grief in cancer care hasn’t distinctly separated it from general grief. What the available evidence does confirm is that the distress HCPs may feel after a patient’s death is a form of grief, while emphasizing its distinctiveness. Professional grief, in contrast to grief occurring over personal losses, involves frequent encounters with patient deaths, though not all patient deaths trigger it due to factors like the depth of the professional relationship. When professional grief does arise, it tends to be less intense, possibly because it’s confined to the workplace. However, this link between grief and the workplace presents unique challenges, including a heightened sense of responsibility and occasional guilt over patient deaths. Additionally, healthcare professionals often perceive their professional grieving reactions as inappropriate or socially unacceptable (see supplementary file 4 for an overview of contrasting elements of professional grief).

While most studies within the corpus of studies reviewed use terms like “grief”, few directly refer to “professional grief” or provide clear definitions. This inconsistent terminology poses a problem, introducing uncertainty in data interpretation. Furthermore, the available quantitative studies have predominantly employed questionnaires tailored to the assessment of grief experienced within personal contexts (40,41). Nonetheless, the distinct nature of grief experienced in a professional context – presumably marked by elements such as guilt stemming from a sense of professional responsibility towards a patient’s life – suggests that conventional assessment tools may fall short in comprehensively capturing professional grief.

### Study limitations

Our scoping review has several limitations. Firstly, our primary search was limited to three electronic databases, which may have caused us to overlook relevant publications. However, we prioritized search sensitivity, evident in the considerable number of abstracts reviewed. Secondly, this review focused solely on professional grief in cancer care. While HCPs in cancer care often face patient deaths, the distinction between them and those in other fields may not accurately reflect reality. It remains uncertain whether the fundamental construct or symptoms differ across disciplines. Furthermore, there was a predominance of certain research groups and geographical locations, limiting generalizability. The absence of insights from the global south and Europe raises doubts about generalizability, despite including papers in English, German, and French. However, our review is the first of its kind to comprehensively address professional grief in cancer care, providing an inclusive overview of the current evidence and valuable insights for future investigations. Another strength is our interdisciplinary author team, including clinical expertise in oncology, palliative care, and psycho-oncology, as well as extensive experience in literature reviews.

### Clinical implications

Despite the absence of a precisely defined construct and the variability in terminology, professional grief undeniably affects HCPs in cancer care. A common theme across reviewed studies is the acknowledgment that witnessing patient deaths may induce emotional distress in HCPs. HCPs express a need for structured guidance on managing patient deaths in cancer care (36). The absence of such guidance may lead to adverse consequences such as compassion fatigue, burnout, and compromised care quality (8,15,18,35). Institutions may be falling short in their responsibility both to their employees and the patients they serve (17,18,26).

### Conclusion

While the exact terminology and clear differentiation remain unclear, the presence of distress is evident and concerning. While some studies have explored contextual factors and effective management of what we might term as “professional grief”, comprehensive investigations are lacking. Future research should prioritize establishing a precise definition of “professional grief” and develop robust measures. Insights should be gathered from various occupations and geographic regions, within and beyond cancer care. These efforts are crucial for advancing towards a healthcare system that prioritizes the well-being of its professionals.

## Supporting information

supplemental file 1, inclusion criteria

supplemental file 2, search strategy

supplemental file 3, definitions

supplemental file 4, contrasting professional grief

## Data Availability

All data produced in the present study are available upon reasonable request to the authors, however given the review nature of this article the amount of produced data is very limited.

## Acknowledgements

We extend our sincere gratitude to our co-authors and our intern, Lisa Weifenbach (LW), for her support during the data extraction of this review.

## Author contribution statement

Isabelle Scholl was the principal investigator of this study. Svenja Wandke and Isabelle Scholl both contributed to the design, conceptualization and preparation of the study.

Data selection, extraction, analysis, and synthesis was primarily conducted by Svenja Wandke, and supported by Mareike Thomas and Hannah Führes. Klaus Lang, Martin Härter and Karin Oechsle advised on conceptualization. All authors contributed to interpretation of results.

Svenja Wandke drafted the manuscript. All authors critically revised the manuscript. The final version was approved by all authors.

