## supplemental file 1, inclusion criteria for "Professional grief in cancer care – A scoping review"

**Checklist Inclusion Criteria**

| ***Publication ID (author, year, title)*** | |
|---|---|
| *General* | Yes/ No |
| **German, English or French language** | ☐ ☐ |
| **Full text available** | ☐ ☐ |
| *Population* | |
| **Health Care Professionals** (e.g. Doctors, Psychologists, Nurses, Social Workers etc.; graduated), professionals involved in medical or paramedical care<br>NOT:<br>in training/ nursing or medicine students/ administrative staff etc. | ☐ ☐ |
| *„Event“* | |
| **A patient’s death**<br>NOT:<br>death of a partner/ friend/ relative or a colleague or supervisor | ☐ ☐ |
| *Concept of interest* | |
| **Professional grief**<br>→ any research pertaining the reaction towards or coping with a patient’s death as one of or the primary outcome(s) | ☐ ☐ |
| *Context* | |
| **Cancer care**<br>→ medical or paramedical (e.g. psychotherapeutical, social work, etc.) care for **patients with cancer** (including palliative and pediatric settings) | ☐ ☐ |
| *Methodology* | |
| **Scientific articles** (Empirical data, original research articles (using qualitative, quantitative, or mixed methods), systematic literature synthesis)<br>NOT:<br>narratives, personal views/ accounts, etc. | ☐ ☐ |
| **References:** | |
| The following references need to be checked: | |
