## supplemental file 2, search strategy for "Professional grief in cancer care – A scoping review"

Supplementary file 1, example for a search strategy

The final search strategy for MEDLINE was the following: ("Health care professionals" OR "medical professionals" OR psychologist OR psychooncologist OR oncologist OR (healthcare provider[MeSH Terms]) OR (health personnel[MeSH Terms])) AND (On-cology OR Cancer OR Psychooncology OR (oncology[MeSH Terms])) AND („patient death" OR "patient loss" OR death) AND (grief[MeSH Terms] OR "professional grief" OR "staff grief" OR (bereavement[MeSH Terms]) OR "professional bereavement" OR grief support OR "grief education" OR coping OR (coping death[MeSH Terms]) OR mourning)
