## supplemental file 3, definitions for "Professional grief in cancer care – A scoping review"

Supplementary file 4, Definitions of professional grief

| Author (year) | Title | Definition of professional grief |
|---|---|---|
| Granek et al. (2012) | Nature and Impact of Grief Over Patient Loss on Oncologists' Personal and Professional Lives. | "In addition to sadness, crying, and loss of sleep, oncologists' grief had unique elements related to their sense of responsibility for their patients' lives. […] Oncologists' grief also included feelings of powerlessness, self-doubt, guilt, and failure." "We found that for oncologists, patient loss was a unique affective experience that had a smoke-like quality. Like smoke, this grief was intangible and invisible. Nonetheless, it was pervasive, sticking to the physicians' clothes when they went home after work and slipping under the doors between patient rooms." |
| Granek et al. (2016) | Barriers and facilitators in coping with patient death in clinical oncology. | "Granek and colleagues found that oncologists' grief was a unique emotional experience that included feelings of self-doubt, guilt, failure, powerlessness, sadness, loss of sleep, and crying." (Referencing Granek et al., 2012) |
| Treggalles & Lowrie (2018) | An exploration of the lived experience of professional grief among occupational therapists working in palliative care settings. | "Grief is the emotional response to the loss of someone or something of value […]. Exposure to sadness and grief is an inevitability for occupational therapists working in palliative care […]. The resultant experience of professional grief among health professionals working in this field is, therefore, neither unexpected nor inappropriate […]." |
