## supplemental file 4, contrasting professional grief for "Professional grief in cancer care – A scoping review"

Supplementary file 5, Contrasting professional grief and grief in general

| **Grief (in general)** | vs. | **Professional Grief** |
|---|---|---|
| Infrequent deaths | | Common (everyday) Deaths |
| Occurs highly likely after each loss | | Does not occur necessarily after every loss (Frequency fluctuations between 23% and 100%) |
| Fundamentally impactful losses | | Losses affect only one area of life |
| High intensity of distress | | Moderate to high intensity of distress |
| No responsibility for the dying process | | Shared responsibility for the dying process, leading to feelings of guilt |
| Perceived as valid grief by mourners | | Perceived as disenfranchised Grief? ("need to 'keep it together'") |
